# A novel homozygous missense mutation p.P388S in *TULP1* causes protein instability and retinitis pigmentosa

**DOI:** 10.1101/2020.12.04.20238931

**Authors:** DaNae R. Woodard, Chao Xing, Pratyusha Ganne, Hanquan Liang, Avinash Mahindrakar, Chandrasekhar Sankurathri, John D. Hulleman, V. Vinod Mootha

## Abstract

**Purpose:** Retinitis pigmentosa (RP) is an inherited retinal disorder that results in the degeneration of photoreceptor cells, ultimately leading to severe visual impairment. We characterized a consanguineous family from Southern India wherein an individual in his 20’s presented with night blindness since childhood. The purpose of this study was to identify the causative mutation for RP in this individual as well as characterize how the mutation may ultimately affect protein function.

**Methods:** We performed a complete ophthalmologic examination of the proband followed by exome sequencing. The identified mutation was then modeled in cultured cells, evaluating its expression, solubility (both by western blot), subcellular distribution (confocal microscopy), and testing whether this variant induced endoplasmic reticulum (ER) stress (qPCR and western blotting).

**Results:** The proband presented with generalized and parafoveal retinal pigment epithelial atrophy with bone spicule pigmentation in the mid periphery and arteriolar attenuation. Optical coherence tomography scans through the macula of both eyes showed atrophy of outer retinal layers with loss of the ellipsoid zone, whereas systemic examination of this individual was normal. The proband’s parents and sibling were asymptomatic and had normal funduscopic examinations. We discovered a novel homozygous p.Pro388Ser mutation in the tubby-like protein 1 (*TULP1*) gene in the individual with RP. In cultured cells, the P388S mutation does not alter the subcellular distribution of TULP1 or induce ER stress when compared to wild-type TULP1, but instead significantly lowers protein stability as indicated by steady-state and cycloheximide-chase experiments.

**Conclusions:** These results add to the list of known *TULP1* mutations associated with RP and suggest a unique pathogenic mechanism in *TULP1*-induced RP, which may be shared amongst select mutations in *TULP1*.

## INTRODUCTION

Inherited retinal degenerations (IRDs) caused by autosomal dominant, recessive, and X-linked mutations comprise more than 2 million cases of ocular diseases worldwide[1]. The most common IRD, retinitis pigmentosa (RP), affects 1 in 3,000 individuals worldwide and is characterized by the degeneration of retinal photoreceptor cells beginning with the atrophy of rods followed by the secondary death of cones[2]. Clinical symptoms of RP include night blindness followed by the loss of peripheral, and eventually, central vision[3].

Currently, more than 30 genes have been associated with autosomal recessive RP[4]. Mutations in the tubby-like protein 1 (*TULP1*) gene have been shown to contribute to the autosomal recessive RP[5-8]. *TULP1* belongs to the tubby-like gene family that encodes for a 542 amino acid cytoplasmic, membrane-associated protein found exclusively in retinal photoreceptor cells[9]. Previously, the TULP1 protein was demonstrated to be required for normal photoreceptor function through promotion of rhodopsin transport and localization from the inner to outer segment[10], potentially in an F-actin-dependent manner[11]. In addition, *in vivo* studies have confirmed that mice lacking *Tulp1* display early-onset photoreceptor degeneration due to the loss of rods and cones[12]. Recently, Lobo et. al. demonstrated that certain RP-associated autosomal recessive missense mutations in the *TULP1* gene can cause the protein to accumulate within the endoplasmic reticulum (ER), leading to prolonged and possibly detrimental ER stress, providing a surprising, but speculative molecular mechanism by which *TULP1* mutations can induce retinal degeneration[13].

In the current study, we identified a novel homozygous missense mutation p.Pro388Sser (P338S) in *TULP1* in a consanguineous family from Southern India that presented with autosomal recessive RP. We explored whether the P388S TULP1 mutant demonstrated any differences in solubility, subcellular localization, or activated cellular stress response. Our observations reveal that there are no differences in transcript levels between P388S and wild-type (WT) *TULP1*, nor does the P388S mutation induce overt ER stress within cells. Furthermore, we found that P388S localized similarly to WT TULP1 in transfected HEK-293A and ARPE-19 cells. However, we did find that P388S steady state levels were significantly reduced and that P388S was more rapidly degraded than WT TULP1 through cycloheximide-chase assays. Our results suggest that particular mutations in *TULP1* may affect protein stability, which may in turn contribute to RP disease pathogenesis.

## METHODS

### Study Subjects

This study was approved by the Institutional Review Board of the Srikiran Institute of Ophthalmology and followed the tenets of the Declaration of Helsinki. The affected proband and family members were recruited and examined after informed consent. All subjects underwent detailed ophthalmologic evaluations including fundus examination by a retina fellowship-trained ophthalmologist.

### Exome Sequencing

We performed exome sequencing on genomic DNA of the proband. Library construction and target enrichment were performed using the IDT xGen Exome capture kit. The libraries were then sequenced to mean 100X on-target depth on an Illumina sequencing platform with 150 base pairs paired-end reads. Sequences were aligned to the human reference genome b37 and variants were called using the Genome Analysis Toolkit[14] and annotated using SnpEff[15].

We filtered for rare missense, nonsense, splicing or frameshift homozygous mutations with the minor allele frequency (MAF) <0.01 in the 1000 Genomes Project (http://www.inter-nationalgenome.org/) and genome aggregation (gnomAD; http://gnomad.broadinstitute.org/) databases. Variants with the Genomic Evolutionary Rate Profiling (GERP^++^) score >2.0 and Combined Annotation Dependent Depletion (CADD) score >15 were considered. Known RP susceptibility conferring genes[16] were screened with priority. Sanger sequencing was utilized to validate variants of interest in the proband and family members.

### Generation of TULP1 Constructs

The cDNA encoding for WT human *TULP1* was purchased from the DNASU Plasmid Repository (HsCD00820883, Tucson, AZ). To generate the P388S mutation, Q5 site-directed mutagenesis (New England Biolabs, NEB, Ipswich, MA) of full-length human *TULP1* was performed using the following primers: 5’-CGGGCAGAACTCACAGCGTGG-3’ and 5’- TTGTCAAAGACCGTGAAGCGG-3’. To generate the C-terminal green fluorescent protein (GFP)-tagged WT and P388S *TULP1*, Gibson Assembly (HiFi Master Mix, NEB) was used to insert a Kozak sequence (DNA sequence: GCCACC) upstream of the *TULP1* start codon, and a flexible linker (amino acids: GGGGS) separating *TULP1* and enhanced GFP (eGFP) into the peGFP-C1 vector backbone via the SalI and NheI restriction sites. All constructs were verified by Sanger sequencing.

### Cell Culture

Human embryonic kidney cells (HEK-293A, Life Technologies, Carlsbad, CA) were cultured at 37°C with 5% CO_2_ in Dulbecco’s minimal essential medium (DMEM) supplemented with high glucose, (4.5 g/L, Corning, Corning, NY), 10% fetal bovine serum (FBS, Omega Scientific, Tarzana, CA) and 1% penicillin-streptomycin-glutamine (Gibco, Waltham, MA). For a 24 well plate, cells were plated at a density of 100,000 cells/well and for a 12 well plate, cells were plated at a density of 180,000-200,000 cells/well. Cells were transfected the following day with either 500 ng (24 well) or 1 μg (12 well) of midi-prepped endotoxin-free plasmid DNA (Qiagen, Germantown, MD) using Lipofectamine 3000 (Life Technologies) according to the manufacturer’s protocol. Forty-eight hours after transfection, fresh media was added and cells were harvested 24 h later (72 h post transfection) and processed for western blot or quantitative PCR (qPCR). As a positive control for some experiments, cells were treated with tunicamycin (an unfolded protein response inducer, 1 μM, 24 h, Sigma) and processed similarly for western blot or qPCR. Human immortalized retinal pigmented epithelial cells (ARPE-19, CRL-2302, American Type Culture Collection, Manassas, VA) were cultured in DMEM/F12 media supplemented with 10% fetal bovine serum (FBS, Omega Scientific, Tarzana, CA), HEPES (Corning, Corning, NY) and penicillin/streptomycin and glutamine (PSQ, Gibco, Germantown, MD). For a 24 well plate, ARPE-19 cells were plated at a density of 100,000 cells/well and transfected the following day with 500 ng of midi-prepped endotoxin-free plasmid DNA (Qiagen, Germantown, MD) using Lipofectamine 3000 (Life Technologies). All cells used were verified for authenticity using short tandem repeat (STR) profiling (Fig. S3, University of Arizona Genomics Core, Tucson, AZ). Note that STR verification cannot distinguish among different variants of the 293-based cell lines (i.e., 293 vs. 293A vs. 293T).

### Confocal Microscopy

A glass bottom 24 well plate (MatTek Corporation) was coated with 1X poly-D-lysine (Sigma Aldrich), rinsed with water and allowed to dry at room temperature. HEK-293A or ARPE-19 cells were plated at a density of 100,000 cells/well and transfected the following day with 500 ng of midi-prepped endotoxin-free plasmid DNA (Qiagen, Germantown, MD). Forty-eight hours after transfection, fresh media was added and 24 h later (72 h post transfection) cells were washed twice with 1X PBS followed by incubation with 4% paraformaldehyde (PFA) (Electron Microscopy Sciences) for 20 minutes. After PFA, cells were washed with 1X PBS. For ARPE-19 cells, cell nuclei were stained with 300 nM 4,6-diamidino-2-phenylindole (DAPI), dilactate solution (Molecular Probes). For membrane staining, HEK-293A cells were washed twice with 1X PBS, fixed in 4% PFA, permeabilized in 0.1% Triton X-100 for 3 min, and washed again in 1X PBS. Cells were incubated in blocking buffer (1% BSA in PBS) for 10 minutes followed by Alexa Fluor™ 633 Phalloidin (1:50 dilution in PBS; Molecular Probes, Eugene, OR) for 20 minutes and washed twice with 1X PBS before being imaged using a 63X oil objective on a Leica SP8 confocal microscope.

### Western Blot Analysis

Cells were washed with Hanks buffered salt solution (HBSS, Corning), then lysed with radioimmunoprecipitation assay (RIPA) buffer (Santa Cruz, Dallas, TX) supplemented with Halt protease inhibitor (Pierce, Rockford, IL) and benzonase (Millipore Sigma) for 3-5 min and spun max speed (14,800 rpm) at 4°C for 10 min. The soluble supernatant was collected and protein was quantified via bicinchoninic assay (BCA) assay (Pierce). The insoluble pellet fractions were further washed in HBSS, centrifuged, and the pellet was resuspended in 1X SDS buffer containing 0.83% BME and sonicated (30% amplitude, pulse 10 sec on/off). Thirty µg of soluble supernatant was run on a 4-20% Tris-Gly SDS-PAGE gel (Life Technologies) alongside the equivalent amount of insoluble protein and transferred onto a nitrocellulose membrane using an iBlot2 device (Life Technologies). After probing for total protein transferred using Ponceau S (Sigma), membranes were blocked overnight in Odyssey Blocking Buffer (PBS) (LICOR). Membranes were then probed with either mouse anti-GFP (1:1000; Santa Cruz, cat #sc-9996), mouse anti-glucose-regulated protein 78 (GRP78,1:1000; Santa Cruz, cat #sc-376768), or rabbit anti-β-actin (1:1000; LI-COR, Lincoln, NE, cat# 926-42210). Blots were imaged on an Odyssey CLx and quantified using ImageStudio (both from LI-COR)

### Quantitative PCR

Transfected HEK-293A cells were trypsinized (0.25% Trypsin EDTA, Gibco), quenched with DMEM, and centrifuged at max speed (14,800 rpm) at 4°C for 10 min. Cell pellets were washed with HBSS, centrifuged again, then RNA extraction from cell pellets was performed using the Aurum Total RNA Mini Kit (BioRad). 400 ng RNA was reverse transcribed using qScript cDNA SuperMix (Quanta Bioscience, Beverly, MA, USA) and the cDNA was diluted 5X in DNase/RNase-free water. cDNA was amplified with TaqMan Fast Advanced Master Mix (Thermo Fisher: Applied Biosystems (cat# 4444963) and h*TULP1* (cat# hs00163236_m1), h*HSPA5* (cat# hs00607129_gH), h*DNAJB9* (cat# hs01052402_m1), h*ASNS* (cat# hs04186194_m1), and h*ACTB* (hs01060665_g1) (Thermo Fisher: Applied Biosystems) and quantified using QuantStudio Real-Time PCR Software (Thermo Fisher: Applied Biosystems).

### Cycloheximide Chase Assay

Twenty-four hours after transfection with WT TULP1 eGFP or P388S TULP1 eGFP constructs, HEK-293A cells were treated in 24 well plates with cycloheximide (25 μM) (Alfa Aesar, cat# J66901-03) for 0, 1, 3, 6, and 9 h. Cells were washed with HBSS, harvested at each time point, and then processed for western blotting.. Membranes were probed with mouse anti-GFP (1:1000; Santa Cruz, cat #sc-9996).

## RESULTS

### Proband from a consanguineous family in South India

An individual in his 20’s (Study ID: SIO221) born of a consanguineous marriage from Southern India, an area where we have previously identified unique autosomal recessive mutations linked to eye disease[17], presented with a history of night blindness since childhood. Best-corrected visual acuity (BCVA) in both eyes was 20/60. He had no nystagmus. Intraocular pressure and anterior segment examinations were normal. Fundus examination revealed a fairly symmetric generalized and parafoveal retinal pigment epithelial (RPE) atrophy with bone spicule pigmentation in the mid periphery and arteriolar attenuation (Fig. 1A, B). The optic nerve head was normal in appearance. Fundus autofluorescence revealed a parafoveal ring of hypo-autofluorescence corresponding to the area of RPE atrophy and patchy decrease in autofluorescence throughout the retina in both eyes (Fig. 1C, D). Optical coherence tomography scans through the macula of both eyes showed atrophy of outer retinal layers with loss of the ellipsoid zone (EZ) and a thin epiretinal membrane (Fig. 1E, F). For comparison, an age-matched healthy control patient was imaged using the same modalities (Fig. 1G-L). There was no evidence of posterior staphyloma in the patient. Patient axial lengths were 24.47 mm and 24.25 mm respectively. Systemic examination was normal. Examined parents and sibling (pedigree shown in Fig. 2A) were asymptomatic and had normal funduscopic examinations.

**Figure 1.**
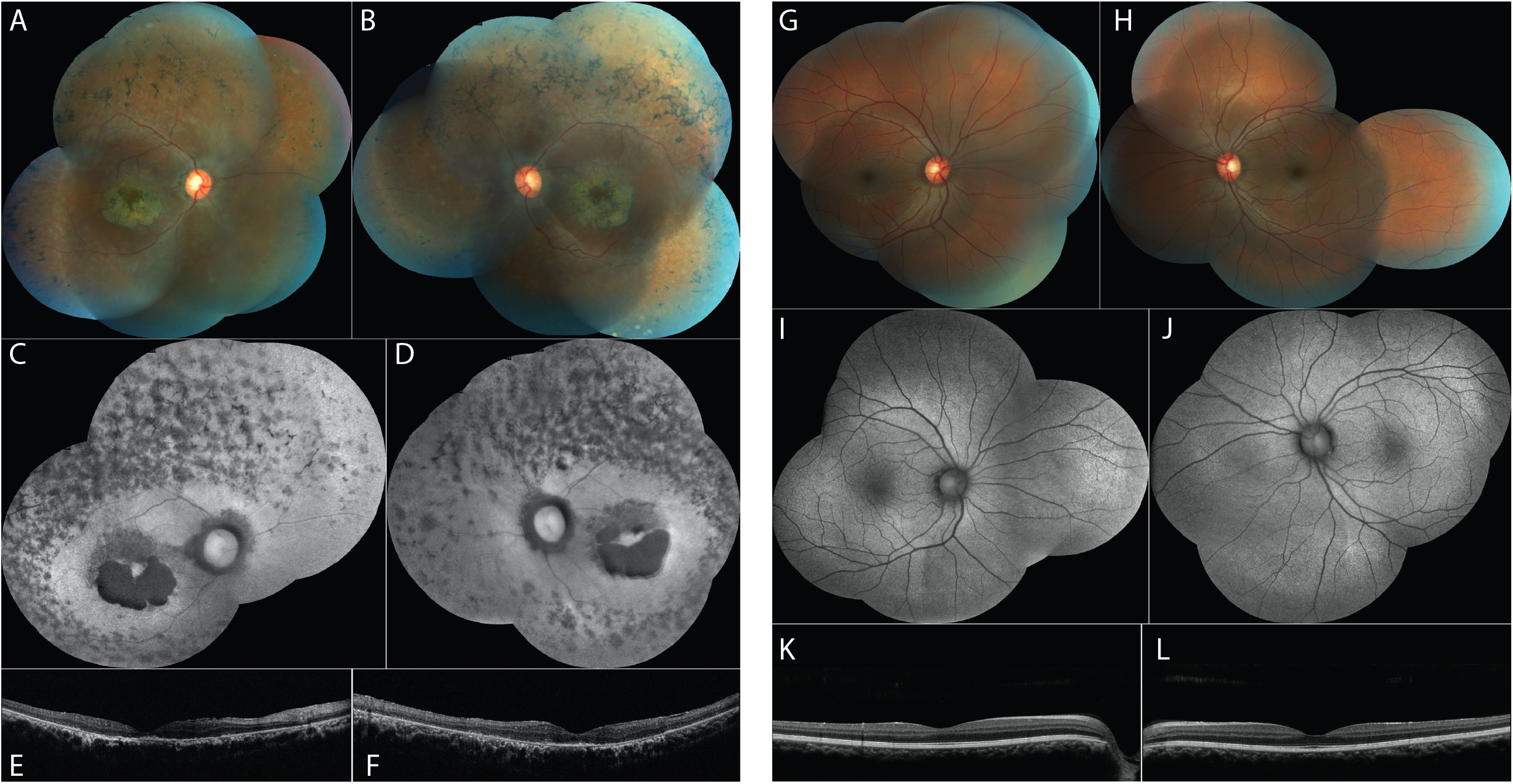
Clinical characterization of the patient. (*A, B*) Fundus photographs of the patient’s right and left eyes showing parafoveal retinal pigment epithelial atrophy, bone spicule pigmentation and arteriolar attenuation. (*C, D*) Fundus autofluorescence images showing parafoveal hypo-autofluorescence corresponding to the area of RPE atrophy and patchy decrease in autofluorescence through-out the retina in both eyes. (*E, F*) OCT scans through the macula showing outer retinal atrophy with loss of the ellipsoid zone. (*G-L*) Fundus photographs, autofluorescence and OCT images of an age-matched control subject

**Figure 2.**
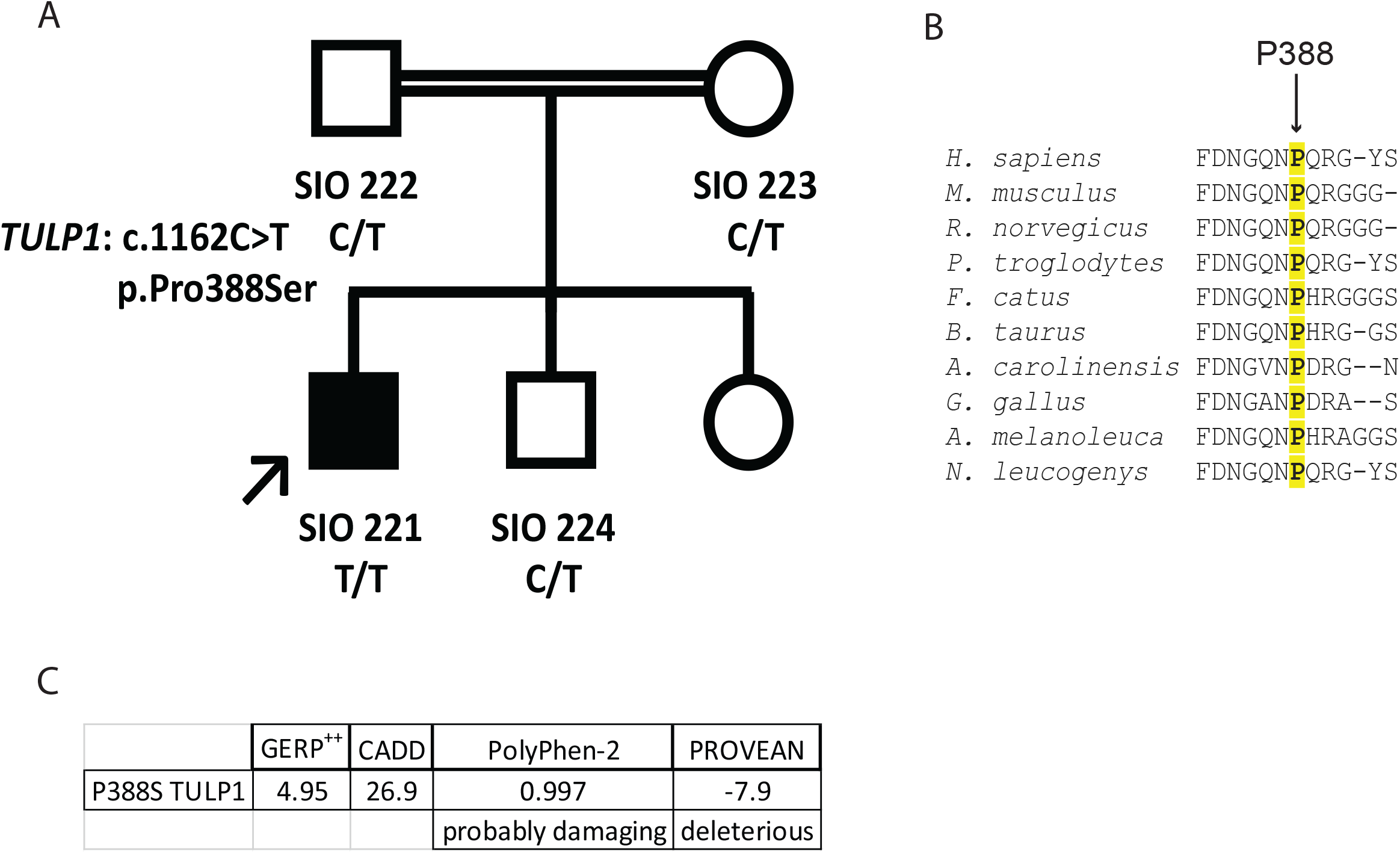
Pedigree and *in silico* analysis of pathogenic mutation (*A*) Pedigree of consanguineous family with variant segregation based on Sanger sequencing. (*B*) Multiple Sequence Alignment of TULP1 amino acid residues across species. Arrow indicates highlighted TULP1 residue. Alignments were performed using Clustal Omega Multiple Sequence Alignment software. (*C*) *In silico* prediction findings related to the P388S mutation.

### Exome sequencing identifies a novel homozygous mutation in *TULP1* gene

Exome sequencing of the proband, followed by application of filtering criteria (described in Methods and the flowchart provided in Fig. S1), revealed ten possible homozygous mutations (Table S1), only one of which was in a gene (*TULP1*) known to cause RP[16]. This variant, a homozygous missense mutation (NC_000006:g.35471576G>A; NM_003322:c.1162C>T; NP_003313:p.Pro388Ser) in exon 12 of the *TULP1* gene, results in a substitution of proline by serine in a conserved amino acid position (Fig. 2B). Aside from the potentially pathogenic *TULP1* mutation, the only known pathogenic mutation (p.Arg89His) that was identified in the affected individual was in the *INS* gene (Table S2), which is associated with hyperproinsulinemia, a disease not known to result in the described ocular phenotype[18]. Nonetheless, the P388S *TULP1* mutation is a novel variant absent from the 1000 Genomes Project database, the Genome Aggregation Database (v2.1.1), the TOPMed database (freeze 5), and the GenomeAsia 100K Project database[19]. Segregation of the variant in the consanguineous pedigree was examined by Sanger sequencing to reveal that the parents are heterozygous for the mutation (Fig. 2A, Fig. S2). P388 is a highly conserved residue amongst the species tested, including mammals (Fig. 2B) with a GERP^++^ score[20] of 4.95 (Fig. 2C). *In silico* prediction indicates that the change to proline at this position could possibly perturb protein function or contribute to pathogenicity with the PolyPhen-2 score[6] of 0.997 (probably damaging), CADD score[21] of 26.9, and a PROVEAN score[22] of -7.9 (deleterious). Analysis of known mutations in *TULP1* shows an enrichment of mutations occurring in the C-terminus of TULP1 (> amino acid 300), with P338S falling within this region (Table S3).

### P388S displays similar subcellular localization to WT TULP1

Previously, WT TULP1 has been shown to localize near the plasma membrane and in the nuclear compartments of COS-7 cells[23]. A separate study suggested that missense mutations in TULP1 shift the sub-cellular trafficking, resulting in ER localization[13]. Therefore, we tested whether the P388S mutant displayed localization differences compared to WT TULP1 protein in cultured cells. HEK-293A cells (STR verified, Fig. S3) were transiently transfected with either eGFP, WT TULP1 eGFP, or P388S TULP1 eGFP tagged constructs and analyzed for green fluorescence and counterstained with phalloidin, which binds to F-actin, using laser-scanning confocal microscopy (Fig. 3A-C). As expected, expression of eGFP showed fluorescent signal distributed evenly across the cytoplasm in cells (Fig. 3A). WT TULP1 eGFP was localized near the plasma membrane as well as nuclear compartments of cells (Fig. 3B) similar to previous reports of TULP1 protein in COS-7 cells[11, 23]. Surprisingly, we found that localization of P388S TULP1 eGFP was not different from that of WT TULP1. P388S TULP1 eGFP was also predominantly localized near the plasma membrane as well as in the nuclear compartment of cells (Fig. 3C), suggesting that there are no differences in cellular distribution between WT and P388S. To confirm that our observations were not cell type-dependent, we also transfected human retinal pigment epithelium (ARPE-19) cells (also STR verified, Fig. S3) with the constructs indicated above and observed that P388S TULP1 eGFP again localized similarly to WT TULP1 eGFP in the nucleus and near the plasma membrane of cells (Fig. S4A-C).

**Figure 3.**
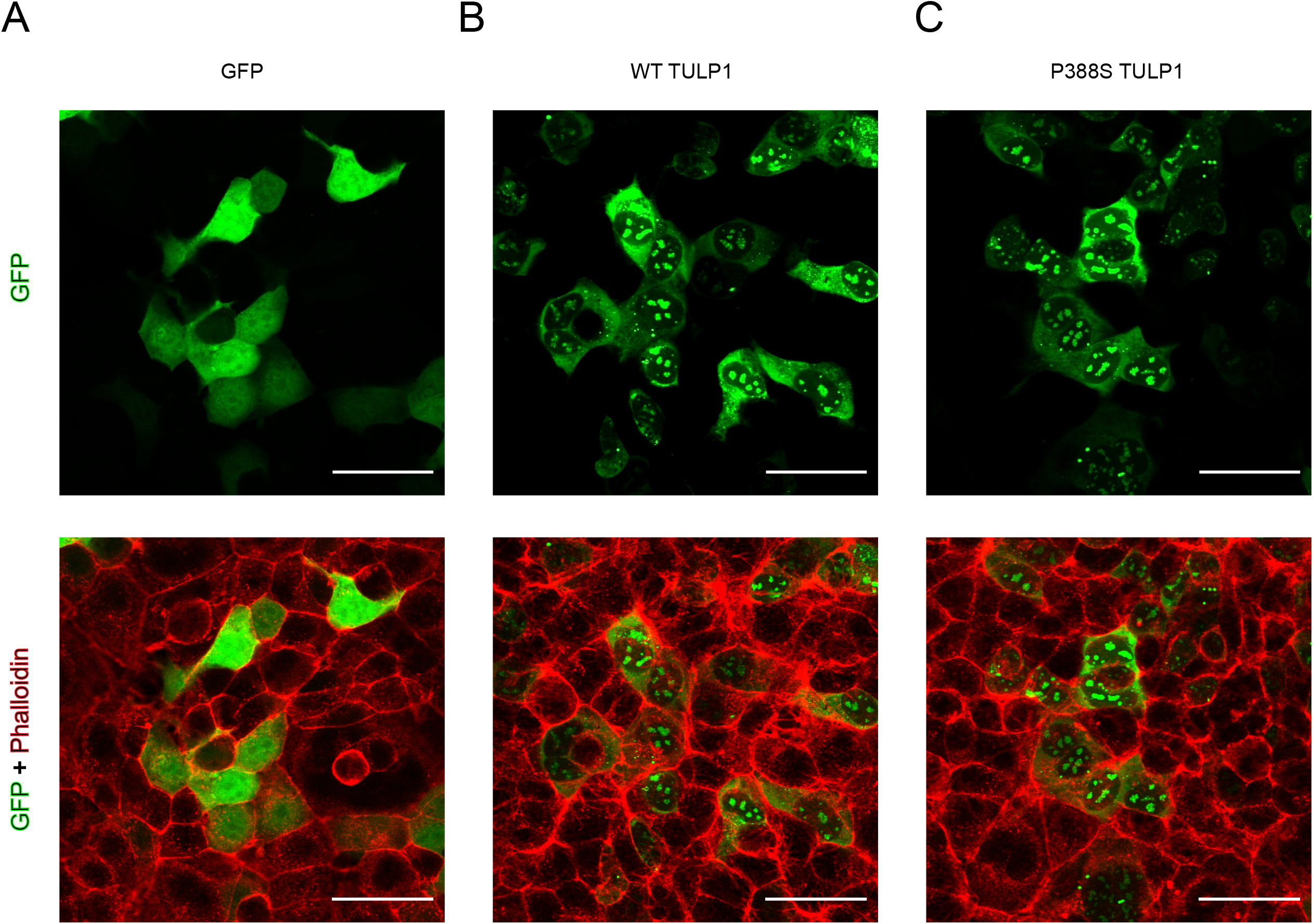
Sub-cellular localization of WT TULP1 and P388S TULP1. Representative confocal microscopy images of HEK-293A cells transfected with (*A*) peGFP-C1, (*B*) WT TULP1 eGFP, or (*C*) P388S TULP1 eGFP constructs (green) and stained with AlexaFluor 633 phalloidin (red).. Scale bar = 50μm. TULP1 eGFP images are representative n ≥ 5 biological, independent replicates. Phalloidin images were representative of n ≥ 3 separate independent wells of a single transfection experiment.

### Protein Expression and Solubility of P388S TULP1

Because we did not detect obvious differences between WT and P388S at the sub-cellular level, we investigated other potential biochemical differences which might partially explain the RP phenotype observed in the patient with the presumed pathogenic variant, p.Pro388Sser in *TULP1*. We employed a biochemical approach to detect expression and solubility of WT and P388S. Using HEK-293A cells, we transfected WT *TULP1* eGFP and P388S *TULP1* eGFP and then 24 h later isolated the soluble and insoluble protein fractions from cells. Both WT TULP1 eGFP and P388S TULP1 eGFP in the soluble and insoluble fractions migrated as predicted at a molecular weight of ∼100kDa (Fig. 4A, TULP1 protein is ∼70kDa[12] and eGFP is ∼26- 28kDa[24]). Both WT TULP1 and P388S TULP1 proteins were similarly more abundant in the RIPA-soluble fraction, as expected based on previous findings[12] (Fig. 4A). However, we detected a significant 27.7 ± 13.8% and 22.2 ± 12.4% decrease in soluble and insoluble P388S TULP1 protein levels compared to WT TULP1, respectively (Fig. 4B). Furthermore, these observed differences were not due to variations at the transcript level since qPCR analysis revealed no significant difference between WT and P388S *TULP1* (Fig. 4C).

**Figure 4.**
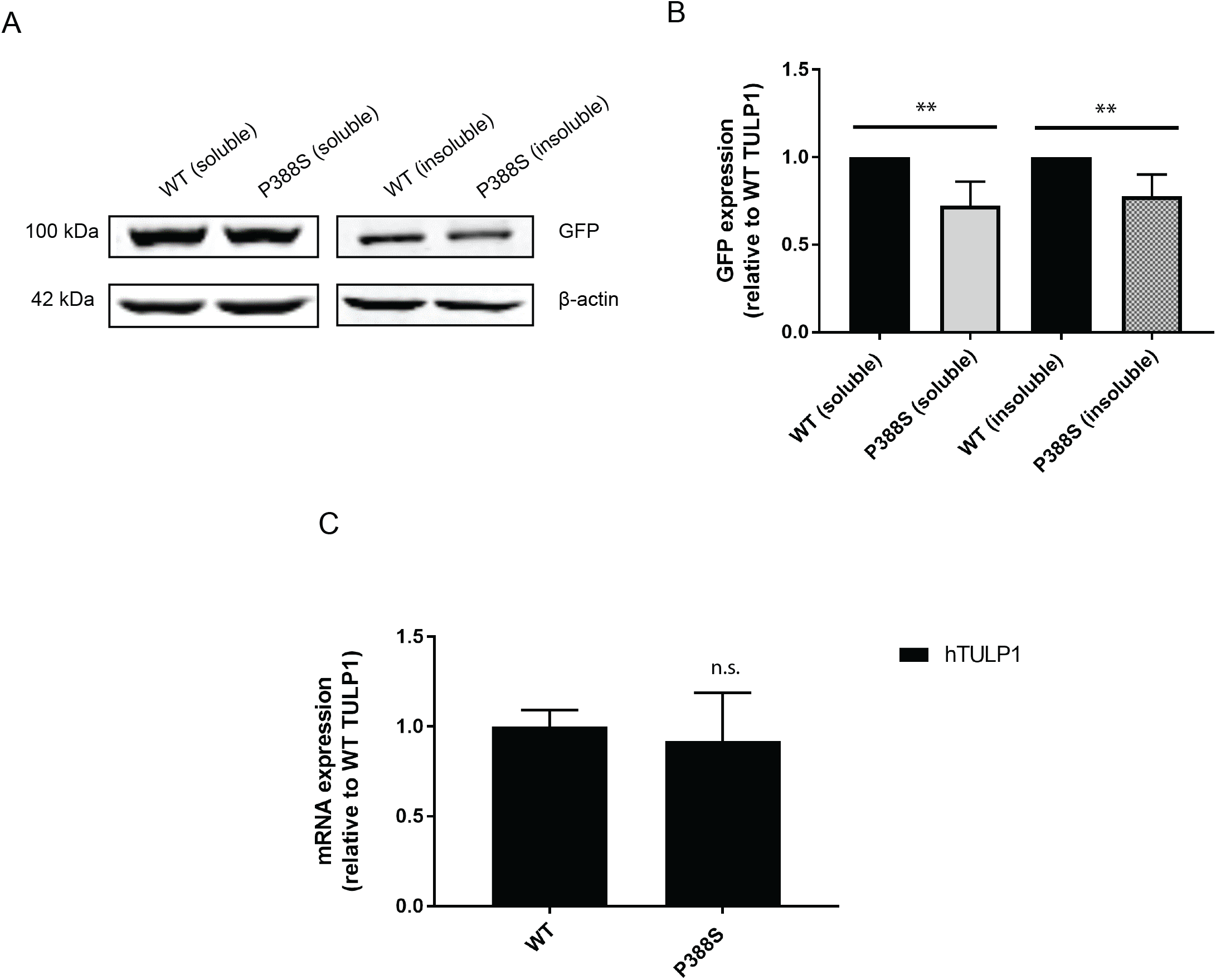
Characterization of the P388S TULP1 variant. (*A*) Western blot of WT and P388S TULP1 eGFP levels in soluble and insoluble fractions. (*B*) Quantification of WT and P388S TULP1 eGFP expression in soluble and insoluble fractions of western blot in (*A*), n ≥ 5, mean ± S.D. (** - p<0.01, one sample t-test vs. hypothetical value of 1 [i.e., unchanged]). (*C*) qPCR analysis of *TULP1* mRNA expression from WT *TULP1* eGFP- and P388S *TULP1* eGFP-transfected HEK-293A cells. Representative data of n ≥ 3 independent experiments, mean ± S.D., n.s. – not significant.

### P388S is degraded more rapidly than WT TULP1

Because we observed a significant reduction in P388S TULP1 protein steady state levels compared to WT TULP1 (Fig. 3A,B), we hypothesized that this may indicate that P388S TULP1 protein is less stable *in vitro*. To more definitively address if there were any differences in stability at the protein level between WT TULP1 and P388S TULP1, transfected cells were treated with cycloheximide (CHX), a translation elongation inhibitor, over the course of 9 h. By western blot, we observed a gradual decrease in protein levels for WT and P388S TULP1 under CHX treatment over time (Fig. 5A, B). Initially, we observed an 18.4 ± 18.2% reduction in P388S levels, compared to 4.8 ± 5.5% reduction of WT TULP1 after 1 h of treatment with CHX (25μM, Fig. 5A-C, not significant). After 3 h of CHX treatment, we observed a significant 48.7 ± 7.9% reduction of P388S levels, in contrast to the stability of WT TULP1 (5.0 ± 12.3%, Fig. 5A-C, p < 0.01, t-test), indicating that P388S is more rapidly degraded at this timepoint. Finally, at 9 hr, we detected a 74.1 ± 11.6% reduction of P388S whereas WT TULP1 only displayed a 53.6 ± 3.1% reduction in protein levels (Fig. 5A-C, p < 0.05, t-test). These data suggest that P388S is generally more unstable and has a higher turnover rate compared to WT TULP1.

**Figure 5.**
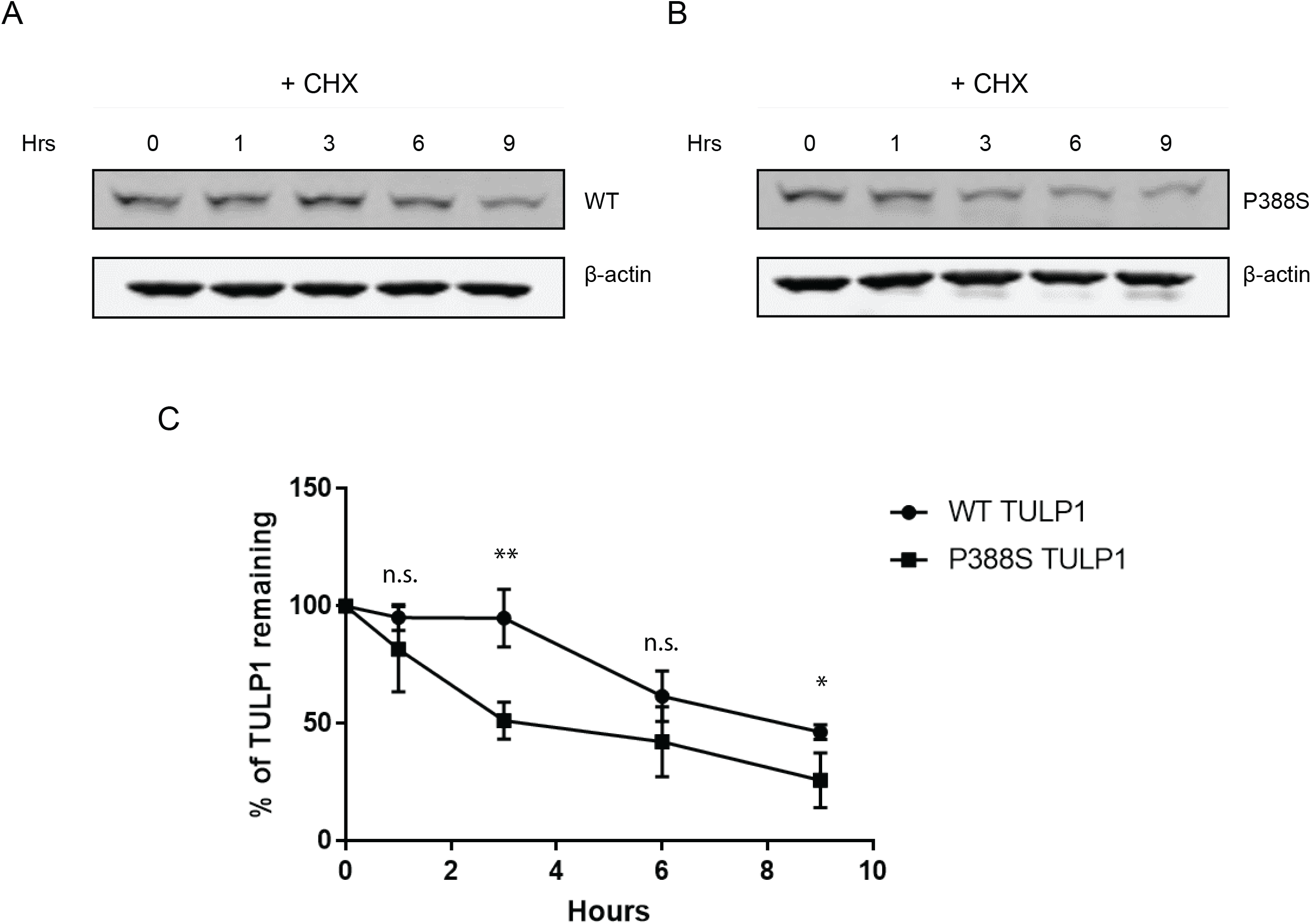
Cycloheximide chase of WT and P388S TULP1. (*A,B*) Western blots of WT and P388S TULP1 eGFP stability in HEK-293A cells treated with 25 μM CHX and harvested at the indicated time points. (*C*) Quantification of western blot from (*A*) and (*B*) showing percentage of TULP1 remaining over time when treated with CHX. (•) indicates WT TULP1 eGFP and (▀) indicates P388S TULP1 eGFP. n = 3 independent experiments, mean ± S.D., * - p < 0.05, ** - p < 0.01, two tailed t-test compared to each WT value, n.s. – not significant.

### P388S TULP1 does not induce ER stress

Missense mutations in *TULP1* have been shown to induce ER stress *in vitro*[13]. Likewise, we hypothesized that P388S TULP1 protein may also induce ER stress in cells. To test this, we transfected HEK-293A cells and performed qPCR using TaqMan probes that are representative downstream genes of unfolded protein response (UPR) pathway activation[25]. To measure changes in ER stress, we selected heat shock protein 70 family protein 5 (*HSPA5*, ATF6 activation), DnaJ homolog subfamily B member 9 (*DNAJB9*, IRE1 activation), and asparagine synthetase (*ASNS*, PERK activation) genes. We measured the mRNA expression levels of each in HEK293A cells expressing either WT or P388S TULP1 and detected no significant differences in *HSPA5, DNAJB9*, and *ASNS* transcript levels (Fig. 6A), suggesting that the presence of P388S does not induce ER stress within cells. We also confirmed these observations at the protein level by analyzing GRP78 (a.k.a. *HSPA5*) levels (Fig. 6B, C). We found that P388S did not induce significant cellular stress in cultured cells when compared to WT TULP1. These results suggest that the P388S TULP1 variant likely contributes to RP by an alternate mechanism other than ER stress.

**Figure 6.**
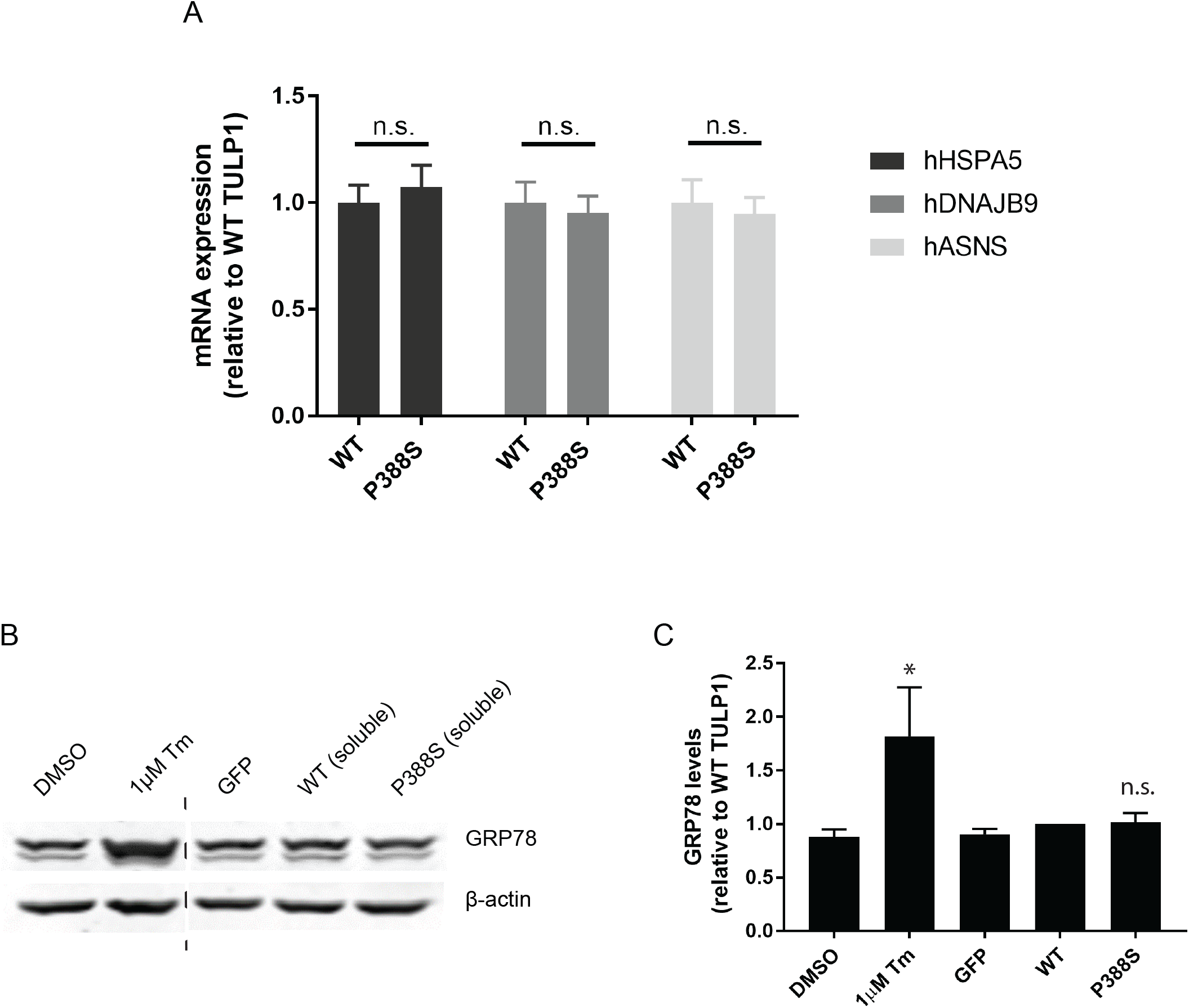
P388S TULP1 does not activate the ER stress response. (*A*) qPCR analysis of h*HSPA5*, h*DNAJB9*, and h*ASNS* transcript levels with TaqMan probes in cells producing WT TULP1 eGFP- or P388S TULP1 eGFP. (*B*) Western blot showing GRP78 expression in eGFP-, WT TULP1 eGFP- or P388S TULP1 eGFP-transfected cells. 1μg/mL Tm was used as a positive control to analyze GRP78 induction. (*C*) Quantification of western blot in (*B*). n=3 biological independent experiments, mean ± S.D., * - p < 0.05, ** - p<0.01, one sample t-test vs. hypothetical value of 1 (i.e., unchanged), n.s. – not significant.

## DISCUSSION

Over twenty-five mutations in *TULP1* have been implicated in RP and Leber’s congenital amaurosis (LCA) including splice-site mutations, frame shift mutations, nonsense mutation and missense mutations[8, 26-32] (Table S3). In the present study, we characterized the P388S TULP1 variant found in an individual with autosomal recessive RP.

When monitoring TULP1 sub-cellular localization in HEK-293A and ARPE-19 cells, as well as ER stress markers as a consequence of *TULP1* expression, we found no obvious differences between WT TULP1 or P388S TULP1-expressing cells. These observations are in contrast to a previous report which showed that missense mutations in *TULP1* can induce ER stress in cultured cells[13]. Our study suggests that not all *TULP1* mutations induce cellular stress that could potentially lead to disease. In cultured HEK-293A cells, we show that in comparison to WT TULP1, the P388S mutant protein is unstable and has a faster turnover. Additional RP-associated mutations in *TULP1* (R311Q and R342Q) were also speculated to cause destabilization of the protein in separate studies [33]. Furthermore, upon closer examination of previous data[13], while not specifically elaborated upon in that particular publication, two other *TULP1* mutations, I459K and F491L, also appear to show a ≥ 45% reduction in apparent steady state levels relative to WT TULP1. While largely speculative, the culmination of these results suggest that a reduction in protein stability might be a phenomenon shared among particular *TULP1* variants.

While the extent of reduction in protein stability/steady-state levels (on average, ∼25%) may not fully explain how the P388S *TULP1* mutation causes RP, this observation indicates that the protein is likely partially misfolded and may be non-functional. To address this possibility, an ideal experiment would be to introduce P388S TULP1 into *Tulp1*^-/-^ mice, to determine whether it can compensate for the loss of *Tulp1*, which is beyond the scope of this study. Nonetheless, our findings suggest the possibility of another avenue, besides ER stress, by which select *TULP1* mutations may lead to disease, and support the idea that evaluation of TULP1 protein stability should be considered when characterizing newly identified *TULP1* mutations associated with RP *in vitro*.

## Supporting information

All supplemental info

## Data Availability

Data in this paper are available upon request and once peer review has been completed.

## ACKOWLEDGEMENTS

DRW is supported by an NIH Diversity Supplement (EY027785-03S). JDH is supported by an endowment from the Roger and Dorothy Hirl Research Fund, an NEI R01 grant (EY027785), and a Career Development Award from Research to Prevent Blindness (RPB). Additional support was provided by an NEI Visual Science Core grant (P30 EY030413) and an unrestricted grant from RPB (both to the UT Southwestern Department of Ophthalmology).

## FIGURE LEGENDS

**Figure S1**. Flowchart of exome sequencing parameters used to identify pathogenic recessive mutations in RP.

**Table S1**. Ten genes identified in the proband were found to be in accordance with autosomal recessive inheritance. Of these genes, *TULP1* was identified as the only RP-associated potentially pathogenic gene.

**Table S2**. Identification of the Arg89His known pathogenic variant in the *INS* gene in the proband.

**Table S3**. Known mutations in *TULP1* identified in patients with RP or LCA.

**Figure S2**. DNA sequencing chromatogram analysis of *TULP1* variant in proband and family members.

**Figure S3**. Demonstration of STR verification of the 293A and ARPE-19 cell lines. Note that STR verification cannot distinguish among different variants of the 293-based cell lines (i.e., 293 vs. 293A vs. 293T).

**Figure S4**. Sub-cellular localization of WT TULP1 and P388S TULP1 in ARPE-19 cells. Representative confocal microscopy images of ARPE-19 cells transfected with (*A*) peGFP-C1, (*B*) WT *TULP1* eGFP, or (*C*) P388S *TULP1* eGFP constructs. GFP signal is detected in cytoplasm and/or nucleus. The nuclei were stained with 4,6-diamidino-2-phenylindole, dilactate (DAPI; blue). Scale bar = 20μm. Representative images from n = 4 biological, independent replicates.

## Notes

### Competing Interest Statement

The authors have declared no competing interest.

