## Supplementary material for "A novel homozygous missense mutation p.P388S in *TULP1* causes protein instability and retinitis pigmentosa": All supplemental info

Fig. S1

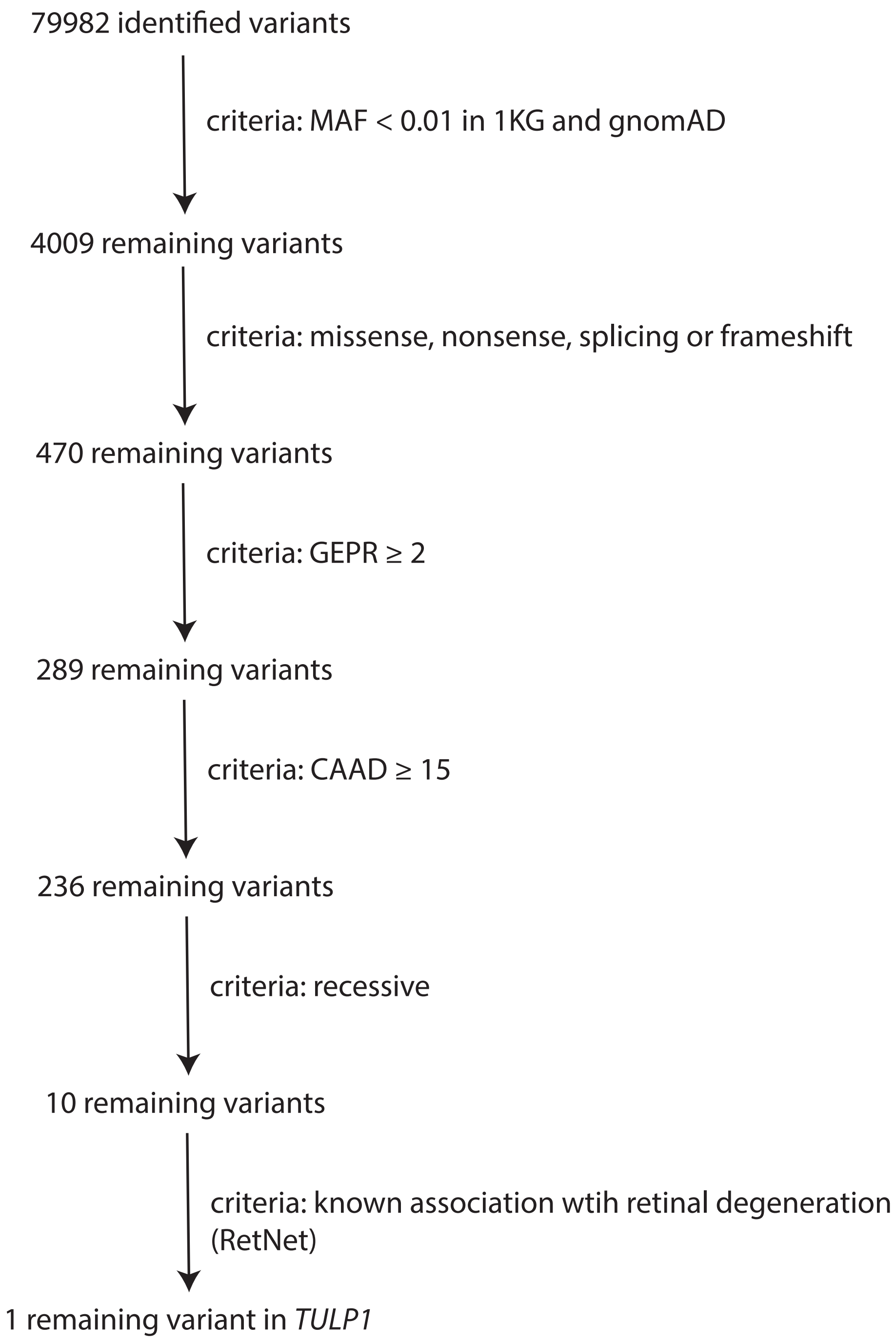
79982 identified variants
criteria: MAF < 0.01 in 1KG and gnomAD
4009 remaining variants
criteria: missense, nonsense, splicing or frameshift
470 remaining variants
criteria: GEPR ≥ 2
289 remaining variants
criteria: CAAD ≥ 15
236 remaining variants
criteria: recessive
10 remaining variants
criteria: known association wtih retinal degeneration (RetNet)
1 remaining variant in *TULP1*

Table S1

| chromosome | position | reference allele | alternative allele | dbSNP ID | variant quality score recalibration | variant type | annotation effect | annotation impact | protein change | gene |
|---|---|---|---|---|---|---|---|---|---|---|
| 1 | 145139088 | C | CG | rs150935736 | 6.18 | INS | splice_acceptor_variant&splice_donor_variant&intron_variant | HIGH | . | NUDT4P1 |
| 12 | 52756723 | G | T | rs745916213 | 3.01 | SNP | missense_variant | MODERATE | p.Thr331Asn | KRT85 |
| 12 | 114384194 | C | G | rs563542363 | 1.25 | SNP | missense_variant | MODERATE | p.Lys498Asn | RBM19 |
| 13 | 73409415 | C | T | rs200956787 | 4.79 | SNP | missense_variant | MODERATE | p.His378Tyr | PIBF1 |
| 13 | 78216919 | G | C | rs770729617 | 3.73 | SNP | missense_variant | MODERATE | p.Glu676Gln | SCEL |
| 6 | 26509316 | C | T | rs781175779 | 1.17 | SNP | missense_variant | MODERATE | p.Pro499Ser | BTN1A1 |
| 6 | 33172447 | T | G | rs552817372 | 7.38 | SNP | start_lost | HIGH | p.Met1? | HSD17B8 |
| 6 | 35471576 | G | A | . | 5.26 | SNP | missense_variant | HIGH | p.Pro388Ser | TULP1 |
| 6 | 43270023 | G | A | rs757872915 | 5.77 | SNP | missense_variant | MODERATE | p.Gly383Arg | SLC22A7 |
| 6 | 51735388 | A | G | rs552199185 | 3.58 | SNP | missense_variant | MODERATE | p.Leu2467Pro | PKHD1 |

### Table S2

| chromosome | position | reference allele | alternative allele | dbSNP ID | variant quality score recalibration | variant type | annotation effect | annotation impact | protein change | gene |
|---|---|---|---|---|---|---|---|---|---|---|
| 11 | 2181149 | C | T | rs28933985 | 3.24 | SNP | missense_variant | MODERATE | p.Arg89His | INS |

Fig. S2

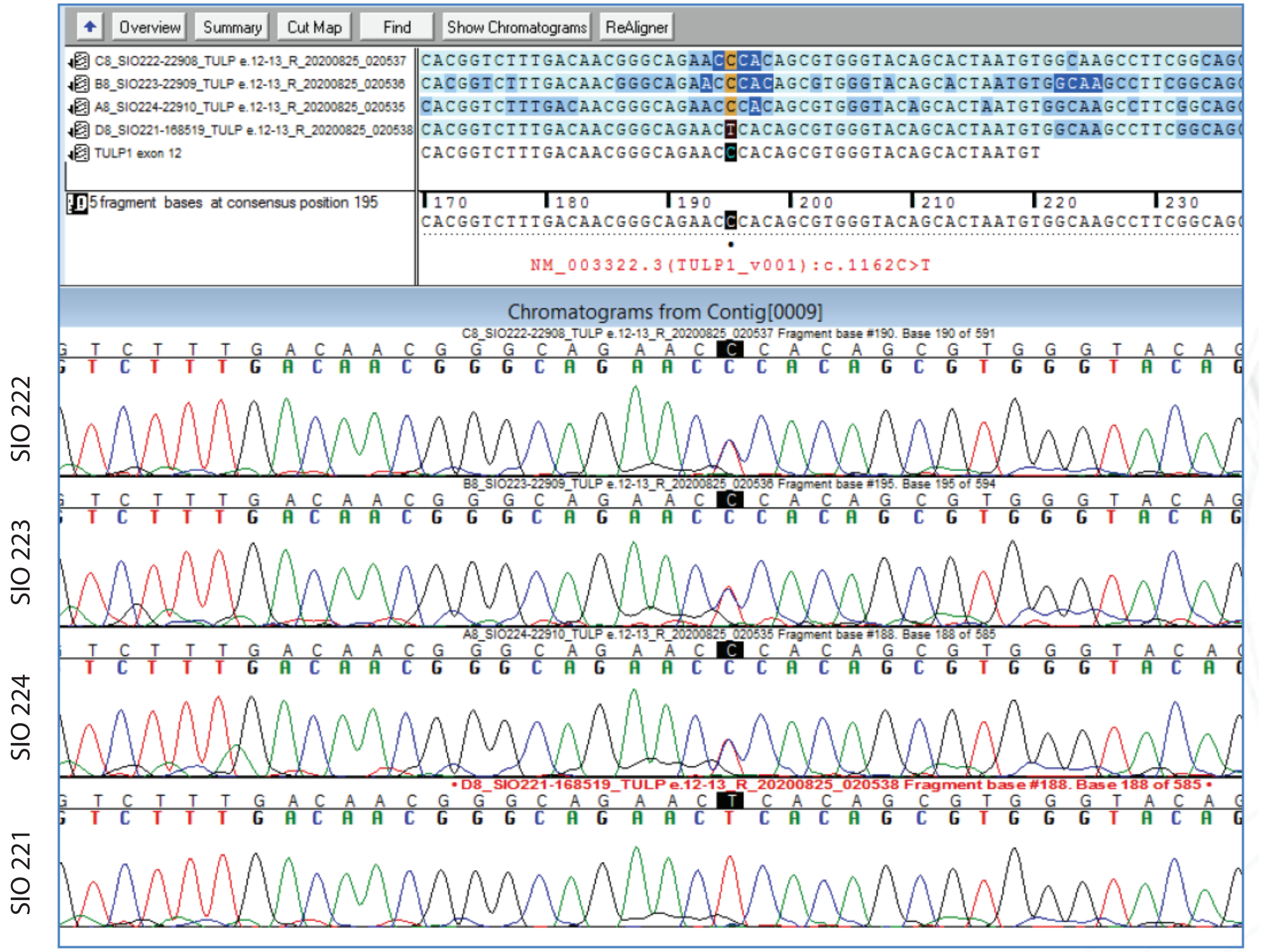

Overview
Summary
Cut Map
Find
Show Chromatograms
ReAligner
C8_SIO222-22908_TULP e.12-13_R_20200825_020537
B8_SIO223-22909_TULP e.12-13_R_20200825_020536
A8_SIO224-22910_TULP e.12-13_R_20200825_020535
D8_SIO221-168519_TULP e.12-13_R_20200825_020538
TULP1 exon 12
5 fragment bases at consensus position 195
NM_003322.3(TULP1_v001):c.1162C>T
Chromatograms from Contig[0009]
C8_SIO222-22908_TULP e.12-13_R_20200825_020537 Fragment base #190. Base 190 of 591
B8_SIO223-22909_TULP e.12-13_R_20200825_020536 Fragment base #195. Base 195 of 594
A8_SIO224-22910_TULP e.12-13_R_20200825_020535 Fragment base #188. Base 188 of 585
D8_SIO221-168519_TULP e.12-13_R_20200825_020538 Fragment base #188. Base 188 of 585
SIO 222
SIO 223
SIO 224
SIO 221

Fig. S3

**Reference STRs**

| Sample Name | D5S818 | D13S317 | D7S820 | D16S539 | VWA | TH01 | AM | TPOX | CSF1PO | Multiple Profiles Seen | Match Comments |
|---|---|---|---|---|---|---|---|---|---|---|---|
| ARPE19-p5 | 13<br>13 | 11<br>12 | 9<br>11 | 9<br>11 | 16<br>19 | 6<br>9.3 | X<br>Y | 9<br>11 | 11<br>11 | NO | Sample matches ARPE-19 above the 80% match threshold. This is considered a match to ARPE-19 (ATCC). |
| HEK-293A-p8 | 8<br>8 | 12<br>14 | 11<br>11 | 9<br>13 | 16<br>19 | 7<br>9.3 | X<br>X | 11<br>11 | 12<br>12 | NO | Sample matches 293T above the 80% match threshold. This is considered a match to 293T (DSMZ). |

**Additional STRs**

| Sample Name | D3S1358 | D21S11 | D18S51 | Penta_E | Penta_D | D8S1179 | FGA |
|---|---|---|---|---|---|---|---|
| ARPE19-p5 | 14<br>15 | 28<br>29 | 12<br>16 | 7<br>11 | 11<br>13 | 13<br>13 | 23<br>23 |
| HEK-293A-p8 | 15<br>17 | 30.2<br>30.2 | 17<br>17 | 7<br>15 | 9<br>9 | 12<br>14 | 23<br>23 |

Fig. S4

ARPE-19

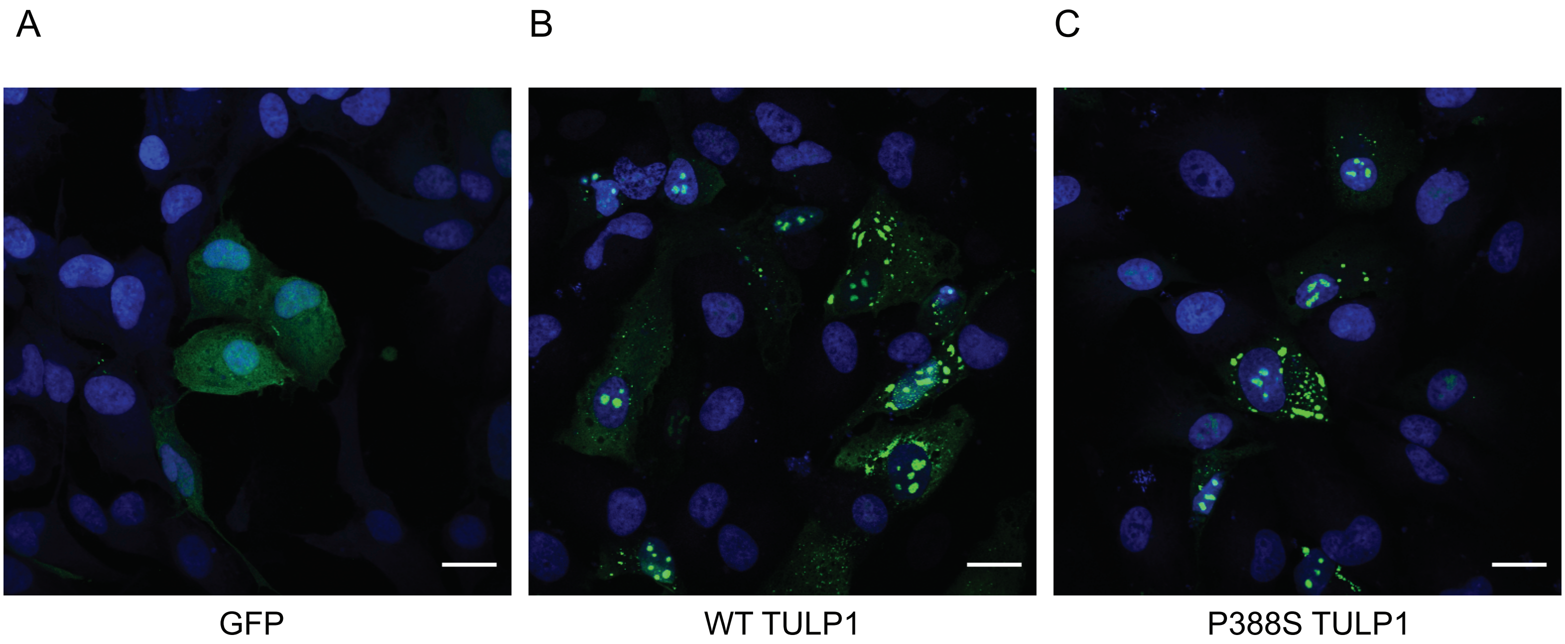

### Table S3

| TULP1 variant | corresponding SNP | associated disease | citiation |
|---|---|---|---|
| D94Y | | LCA | Beryozkin et al (2014) Invest. Ophthalmol. Vis. Sci. |
| S210X | | RP | Glockle et al (2014) Eur. J. Hum. Genet. * |
| A245V | dbSNP:rs62636707 | RP | Uniprot (https://www.uniprot.org/uniprot/O00294) |
| K261T | | RP | Uniprot (https://www.uniprot.org/uniprot/O00294) |
| Q301X | | "TULP1 retinal degeneration" | Li et al (2001) Invest. Ophthalmol. Vis. Sci.; Jacobson et al (2014) Invest. Ophthalmol. Vis. Sci. |
| Q301fsX8 | | early onset RP | Paloma et al (2000) Invest. Ophthalmol. Sci. |
| R311Q | | RP | Hebrand et al (2011) Eur. J. Hum. Genet. |
| R311W/Q492R | | LCA | Tajiguli et al (2016) Sci. Rep. |
| G319D/R482W | | RP | Consugar et al (2015) Genet. Med. |
| Y321D | | LCA | Wang et al (2013) J. Med. Genet. |
| Y321D/R400Q | | "TULP1 retinal degeneration" | Jacobson et al (2014) Invest. Ophthalmol. Vis. Sci. |
| R342Q | | RP | Hebrand et al (2011) Eur. J. Hum. Genet. |
| N349K | | RP | Kannabiran et al (2012) Mol. Vis. |
| D355V | | LCA | Wang et al (2013) J. Med. Genet. |
| D355V/G368W | | "TULP1 retinal degeneration" | Jacobson et al (2014) Invest. Ophthalmol. Vis. Sci. |
| R361X/R420S | | LCA | Glockle et al (2014) Eur. J. Hum. Genet. |
| G363R | | cone/cone-rod dystrophy | Boulanger-Scemama et al (2015) Orphanet J. Rare Dis. |
| G368W | dbSNP:rs387906837 | LCA | Hanein et al (2004) Hum. Mut. |
| R378H | dbSNP:rs148749577 | RP | Uniprot (https://www.uniprot.org/uniprot/O00294) |
| T380A | | LCA | McKibbin et al (2010) Arch. Ophthalmol., Ajmal et al (2012) Mol. Vis. |
| F382S | dbSNP:rs121909076 | RP | Kondo et al (2004) Invest. Ophthalmol. Vis. Sci. |
| G385R | | LCA | Wang et al (2015) Invest. Ophtalmol. Vis. Sci. |
| P388S | | RP | this study |
| R400W | dbSNP:rs387906836 | LCA15/"TULP1 retinal degeneration" | Hanein et al (2004) Hum. Mut.; Jacobson et al (2014) Invest. Ophthalmol. Vis. Sci. |
| R400Q | | RP | Singh et al (2009) Invest. Ophtalmol. Vis. Sci. |
| E402X | | LCA | Hanein et al (2004) Hum. Mut. |
| A405P | | RP | Ge et al (2015) Sci. Rep. |
| R416C | | RP | Katagiri et al (2014) PLoS One |
| R419W | | retinal dystrophy | Sanchez-Alcudia (2014) Invest. Ophthalmol. Vis. Sci. |
| R420P/F491L | dbSNP:rs121909073, dbSNP:rs121909074 | RP | Hagstrom et al (1998) Nat. Genet. |
| R420S | | cone dysfunction | Roosing et al (2013) Ophthalmol. |
| P426L/F506L | | LCA | Wang et al (2013) J. Med. Genet. |
| R440X | | LCA | Wang et al (2015) Invest. Ophtalmol. Vis. Sci. |
| W450X | | LCA/Coat's-like changes | Beryozkin et al (2014) Invest. Ophthalmol. Vis. Sci. |
| T454M | dbSNP:rs138200747 | RP | Hagstrom et al (1998) Nat. Genet. |
| I459K | dbSNP:rs121909075 | RP | Hagstrom et al (1998) Nat. Genet. |
| L461V | | RP | den Hollander et al (2007) Invest. Ophthalmol. Vis. Sci. |
| R482Q | | RP | Ajmal et al (2012) Mol. Vis. |
| R482W/L504fsX140 | dbSNP:rs121909077 | RP | den Hollander et al (2007) Arch. Ophthalmol. |
| Q492R | | LCA | Tajiguli et al (2016) Sci. Rep. |
| P499S | | RP | Ge et al (2015) Sci. Rep. |
| F506L | | LCA | Wang et al (2013) J. Med. Genet. |
| FA531-532 dup. | | LCA/RP | Mataftsi et al (2007) Invest. Ophthalmol. Vis. Sci. |
| F535S | | LCA | Eisenberger et al (2013) PLoS One |
| | | | * = additional mutations identified in USH2A, ABCA4, PRCD identified in this patient |
